# Statistical analysis of sexual dimorphism in adults of Western UP by Computed Tomography based Sternal Morphometry

**DOI:** 10.64898/2026.07.28.26359105

**Authors:** Harsh Kumar Singh, Raushan Kumar, Swati Dixit

## Abstract

**Background:** Sternal dimensions show population-specific sexual dimorphism and may support forensic identification when more informative skeletal elements are unavailable. Computed tomography (CT) permits non-destructive, reproducible three-dimensional assessment of the sternum.

**Objective:** To quantify sex- and age-associated differences in CT-derived sternal measurements among adults from Western Uttar Pradesh, India.

**Methods:** This prospective cross-sectional study included 250 adults aged 20-80 years (155 males and 95 females) referred for chest CT at a tertiary hospital in Moradabad. Images were acquired using a Philips Ingenuity Core 128-slice scanner with 1-mm sections and 0.5-mm reconstruction increments. Manubrium width (MW), manubrium length (ML), sternal body length (B), corpus sterni widths at the first and third sternebrae (CSWS1 and CSWS3), sternal index (SI), combined length (CL), and sternal area (SA) were evaluated. Independent-samples t tests, one-way analysis of variance with Tukey post hoc testing, and Pearson correlations were used.

**Results:** Mean age was 48.1 ± 14.3 years. Males had greater MW (57.3 ± 5.0 vs 49.9 ± 4.0 mm), ML (45.3 ± 5.5 vs 40.4 ± 6.0 mm), B (90.2 ± 9.6 vs 75.7 ± 7.7 mm), CSWS1 (26.1 ± 3.6 vs 23.0 ± 3.3 mm), CSWS3 (31.6 ± 4.9 vs 27.9 ± 4.4 mm), CL (135.5 ± 10.6 vs 116.1 ± 10.0 mm), and SA (5258.3 ± 754.3 vs 3880.5 ± 603.1 mm^2^; all p < 0.001). SI was greater in females (53.8 ± 9.4 vs 50.8 ± 8.5; p = 0.009). Across age groups, only MW (p = 0.001) and SA (p = 0.002) differed significantly. Strong correlations were observed for B with CL (r = 0.902), CL with SA (r = 0.895), B with SA (r = 0.869), and MW with SA (r = 0.847; all p < 0.001).

**Conclusion:** CT-derived sternal morphometry demonstrates marked sexual dimorphism in this Western Uttar Pradesh sample, particularly for SA, CL, B, and MW. These reference values provide a basis for population-specific forensic models; however, predictive accuracy cannot be inferred without classifier development and validation.

## Introduction

Estimation of biological sex is a central component of the forensic biological profile. Although the pelvis and skull usually provide the strongest morphological information, these elements may be absent or damaged in fragmented, decomposed, burned, or mass-fatality remains. The sternum is compact, centrally located, and frequently recoverable; its linear dimensions therefore offer a useful supplementary source of sex-related information [1,2]. Multidetector CT enables thin-section acquisition, multiplanar reconstruction, and three-dimensional visualization without maceration or destructive handling. Previous CT studies in Turkish, Western Australian, Japanese, Saudi, Croatian, Jordanian, Iranian, and other populations have consistently found larger absolute sternal dimensions in males, while the sternal index is often higher in females [3-13]. However, the relative performance of individual measurements and decision thresholds varies across populations. This population dependence is especially relevant in India, where regional genetic, nutritional, environmental, and secular influences may limit direct transfer of standards derived elsewhere. Studies of Indian skeletal and CT samples demonstrate the value of regional reference data and multivariable modelling, but data from Western Uttar Pradesh remain limited [10,14-16]. The present study therefore assessed eight CT-derived sternal parameters in adults from Western Uttar Pradesh. The primary objective was to test whether these measurements differed between males and females. Secondary objectives were to evaluate age-group patterns and correlations among the measurements. It was hypothesised that absolute dimensions and sternal area would be greater in males, whereas the sternal index would show an inverse pattern.

## Materials and Methods

### a) Study design and setting

A clinical, prospective, cross-sectional study was conducted over one year in the Department of Radio-diagnosis and Imaging, Teerthanker Mahaveer Hospital and Research Centre, Moradabad, Uttar Pradesh, India. The source population comprised patients from Western Uttar Pradesh referred from outpatient or inpatient services for a clinically indicated chest CT examination.

### b) Participants

Convenience sampling was used. Adults aged 20-80 years who underwent chest CT and had an adequately visualized sternum were eligible. Exclusion criteria were age below 20 or above 80 years, pathological sternal fusion, previous sternal surgery, previous sternal fracture or malunion, and atypical sternal development. The final sample comprised 250 participants.

### c) CT acquisition and image reconstruction

Imaging was performed using a Philips Ingenuity Core 128-slice multidetector CT scanner. Participants were positioned head first and supine. The acquisition extended from approximately 1 cm above the lung apices to the costophrenic angles in a caudocranial axial helical acquisition. The reported protocol used 120 kVp, 250-300 mAs, 1-mm section thickness, and 0.5-mm reconstruction increment. Multiplanar reformations, maximum-intensity projections, and volume-rendered three-dimensional reconstructions were generated on the Philips Ingenuity workstation.

### d) Sternal measus

Measus were obtained on appropriately aligned multiplanar and 3D volume-rendered images. The primary linear measurements were manubrium width (MW), manubrium length (ML), sternal body length (B), corpus sterni width at the first sternebra (CSWS1), and corpus sterni width at the third sternebra (CSWS3). Derived parameters were combined length (CL = ML + B), sternal index (SI = ML/B × 100), and sternal area [SA = (ML + B) × (MW + CSWS1 + CSWS3)/3], consistent with established CT sternal-morphometry methods [3,11]. Linear measurements were reported in millimeters and area in square millimeters (**fig.1**).

**Figure 1.**
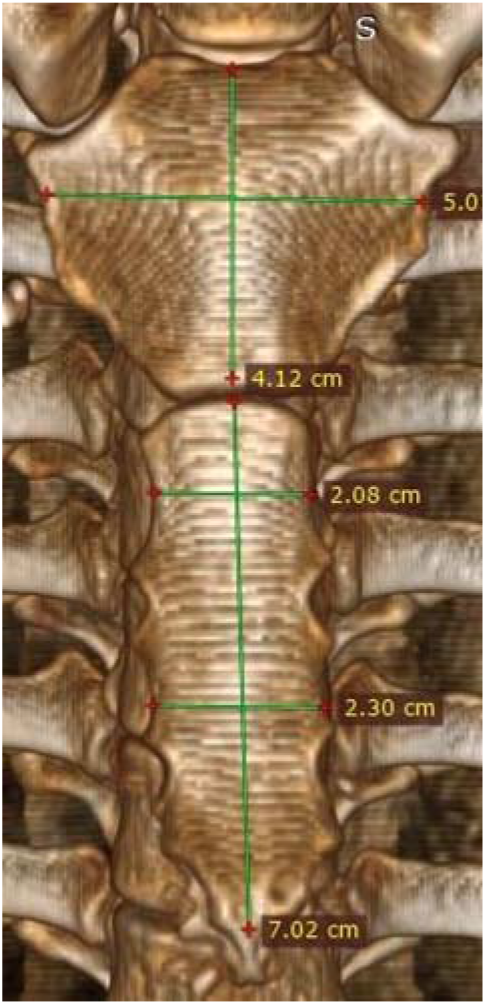
Representative anterior 3D volume-rendered CT image illustrating linear sternal measurements. Values displayed on the source image are in centimeters; analyses were reported in millimeters (area in mm^2^).

### e) Statistical analysis

Analyses were performed using IBM SPSS Statistics version 29.0.10 (SPSS Inc., Chicago, IL, USA, as reported in the thesis). Continuous data were summarized as mean ± standard deviation (SD) and range; categorical data were summarized as frequency and percentage. Male and female measurements were compared using independent-samples t tests. One-way analysis of variance (ANOVA) assessed differences among age groups (20-35, 36-55, and 56-80 years), with Tukey testing for significant pairwise comparisons. Pearson correlation coefficients quantified relationships among sternal measurements. Statistical significance was set at p < 0.05. No discriminant function, logistic-regression classifier, receiver-operating-characteristic analysis, or cross-validation was reported; consequently, the study evaluates dimorphism but does not estimate diagnostic accuracy.

### f) Ethical considerations

The thesis records approval by the Paramedical Research Committee of the College of Paramedical Sciences, Teerthanker Mahaveer University.

## Results

### a) Participant characteristics

The study included 250 participants: 155 males (62.0%) and 95 females (38.0%). Age ranged from 20 to 80 years, with a mean of 48.1 ± 14.3 years. Nearly half of the sample was aged 36-55 years (49.2%). The sex-by-age totals in **Table 1** are internally consistent with the overall sample counts.

**Table 1.** Distribution of participants by age group and sex.

| Age group | Male, n | Male, % | Female, n | Female, % | Total, n (%) |
| --- | --- | --- | --- | --- | --- |
| 20-35 years | 27 | 17.4 | 23 | 24.2 | 50 (20.0) |
| 36-55 years | 73 | 47.1 | 50 | 52.6 | 123 (49.2) |
| 56-80 years | 55 | 35.5 | 22 | 23.2 | 77 (30.8) |
| Total | 155 | 100.0 | 95 | 100.0 | 250 (100.0) |

\*Percentages in the male and female columns are within-sex percentages.

### b) Sex differences in sternal measus

All eight parameters differed significantly between males and females. Seven absolute size measures were greater in males, whereas SI was greater in females. The largest test statistics were observed for SA (t = 15.09), CL (t = 14.35), B (t = 12.51), and MW (t = 12.24), indicating that these variables showed the clearest separation of group means in this sample (**table 2**).

**Table 2.** Comparison of sternal measurements between males and females.

| Measurement | Male (n = 155) | Female (n = 95) | t | p value |
| --- | --- | --- | --- | --- |
| Manubrium width, MW (mm) | 57.3 ± 5.0 | 49.9 ± 4.0 | 12.24 | <0.001 |
| Manubrium length, ML (mm) | 45.3 ± 5.5 | 40.4 ± 6.0 | 6.58 | <0.001 |
| Sternal body length, B (mm) | 90.2 ± 9.6 | 75.7 ± 7.7 | 12.51 | <0.001 |
| Width at first sternebra, CSWS1 (mm) | 26.1 ± 3.6 | 23.0 ± 3.3 | 6.70 | <0.001 |
| Width at third sternebra, CSWS3 (mm) | 31.6 ± 4.9 | 27.9 ± 4.4 | 6.03 | <0.001 |
| Sternal index, SI | 50.8 ± 8.5 | 53.8 ± 9.4 | -2.65 | 0.009 |
| Combined length, CL (mm) | 135.5 ± 10.6 | 116.1 ± 10.0 | 14.35 | <0.001 |
| Sternal area, SA (mm<sup>2</sup>) | 5258.3 ± 754.3 | 3880.5 ± 603.1 | 15.09 | <0.001 |

\*Values are mean ± SD. Independent-samples t test. Negative t for SI reflects a higher female mean.

### c) Age-group patterns

Across the three age groups, significant differences were limited to MW (F = 7.42, p = 0.001) and SA (F = 6.30, p = 0.002). Tukey analysis showed that participants aged 56-80 years had higher MW than those aged 20-35 years (mean difference 3.80 mm, p = 0.001) and 36-55 years (2.39 mm, p = 0.012). SA was likewise higher in the 56-80-year group than in the 20-35-year (mean difference 569.45 mm^2^, p = 0.003) and 36-55-year groups (379.86 mm^2^, p = 0.017). Other measures did not differ significantly by age group (**table 3**).

**Table 3.** Sternal measurements according to age group.

| Measurement | 20-35 years | 36-55 years | 56-80 years | F | P value |
| --- | --- | --- | --- | --- | --- |
| MW (mm) | 52.6 ± 5.0 | 54.0 ± 5.7 | 56.4 ± 6.2 | 7.4<br>2 | 0.001 |
| ML (mm) | 44.2 ± 6.5 | 42.8 ± 6.6 | 43.8 ± 5.2 | 1.1<br>5 | 0.318 |
| B (mm) | 82.8 ± 11.7 | 84.1 ± 11.5 | 86.9 ± 10.6 | 2.2<br>6 | 0.106 |
| CSWS1 (mm) | 24.2 ± 3.3 | 24.8 ± 3.9 | 25.6 ± 4.0 | 2.3<br>4 | 0.098 |
| CSWS3 (mm) | 29.4 ± 4.6 | 30.0 ± 5.1 | 30.9 ± 5.4 | 1.4<br>8 | 0.230 |
| SI | 54.0 ± 8.7 | 51.6 ± 9.4 | 51.1 ± 8.3 | 1.7<br>5 | 0.176 |
| CL (mm) | 127.0 ± 15.1 | 126.9 ± 14.5 | 130.7 ± 12.2 | 1.9<br>1 | 0.150 |
| SA (mm<sup>2</sup>) | 4466.0 ± 912.2 | 4655.6 ± 958.3 | 5035.5 ± 956.1 | 6.3<br>0 | 0.002 |

\*Values are mean ± SD. F statistics are from one-way ANOVA.

### d) Correlations among sternal measurements

The strongest positive relationships reflected shared variation in sternal size. B was strongly associated with CL and SA, while CL and SA were also strongly correlated. SI had an inverse relationship with B, CL, and SA, but was not significantly correlated with MW, CSWS1, or CSWS3. These results describes the B-SI relationship as positive despite the reported matrix showing r = -0.588 (**table 4**).

**Table 4.** Selected Pearson correlations among sternal measurements.

| Variable pair | r | p value | Interpretation |
| --- | --- | --- | --- |
| B with CL | 0.902 | <0.001 | Strong positive |
| CL with SA | 0.895 | <0.001 | Strong positive |
| B with SA | 0.869 | <0.001 | Strong positive |
| MW with SA | 0.847 | <0.001 | Strong positive |
| CSWS1 with CSWS3 | 0.695 | <0.001 | Positive |
| ML with SI | 0.653 | <0.001 | Positive |
| B with SI | -0.588 | <0.001 | Inverse |
| SI with SA | -0.304 | <0.001 | Inverse |
| SI with CL | -0.188 | 0.003 | Weak inverse |
| MW with SI | 0.008 | 0.904 | Not significant |
| CSWS1 with SI | 0.044 | 0.486 | Not significant |
| CSWS3 with SI | 0.008 | 0.897 | Not significant |

\*Pearson correlation coefficients calculated across all 250 participants.

## Discussion and forensic implications

This study demonstrates broad sexual dimorphism in CT-derived sternal dimensions among adults from Western Uttar Pradesh. Males had significantly greater mean values for MW, ML, B, CSWS1, CSWS3, CL, and SA, whereas females had a modestly higher SI. The largest between-sex test statistics occurred for SA, CL, B, and MW. These findings support the hypothesis that both longitudinal and transverse growth contribute to sex-related variation in the adult sternum. The observed pattern is consistent with CT studies in several populations. Kalbouneh et al. found SA, B, CL, and MW among the most dimorphic variables in a large Jordanian sample [11]. Franklin et al. reported that sternal measurements could support sex estimation in Western Australian adults, while emphasising population-specific functions [4]. Studies in Turkish [3,5,8,13], Saudi [7], Japanese [6], Croatian [9], Iranian [12], Sudanese [17], and Egyptian [18] samples have similarly identified significant sex differences in absolute sternal dimensions. The direction of the SI result also agrees with reports that a shorter manubrium relative to body length characterises male sterna in many groups [3,5]. The magnitude and ranking of variables nevertheless differ across studies. In the present sample, SA and CL produced the largest t statistics, followed by B and MW. Because SA combines longitudinal and transverse dimensions, it may capture overall sternal size more comprehensively than a single linear measure. The strong correlations of SA with MW, B, and CL are therefore expected, but also indicate substantial collinearity. Any future multivariable classifier should assess variance inflation, model parsimony, calibration, and overfitting rather than automatically including every correlated measurement.

Recent Indian evidence reinforces the need for regional standards. Dry-bone studies from Northwest India and Bengal have reported population-specific dimorphism and classification rules [14,15]. An Indian MDCT study used discriminant and logistic models to classify sex [10], and a 2025 Central Indian study further demonstrated the value of CT-derived sternal morphometry [16]. The present Western Uttar Pradesh data add a distinct regional sample, but the mean comparisons should not be treated as a ready-to-use identification rule. Only MW and SA varied significantly across age groups, with the highest values in participants aged 56-80 years. This may reflect cohort effects, continuing remodelling, sex composition, or residual differences in body size rather than simple age-related growth. Notably, the older group contained a larger proportion of males than the youngest group. Age and sex should therefore be modelled simultaneously in future analyses using the individual-level dataset.

The findings identify SA, CL, B, and MW as priority variables for developing a Western Uttar Pradesh sex-estimation model. CT offers a non-destructive route for building contemporary population databases from living or postmortem imaging and can be especially useful when the sternum is intact but the pelvis or skull is unavailable. Before operational use, however, thresholds or equations must be derived from the raw data, internally validated by resampling or cross-validation, tested in an independent regional sample, and accompanied by uncertainty estimates.

## Conclusion

Adults from Western Uttar Pradesh showed significant sex-related differences in all eight evaluated sternal parameters. Absolute dimensions and area were greater in males, while the sternal index was greater in females. Sternal area, combined length, body length, and manubrium width produced the clearest group differences and are strong candidates for future model development. These data establish regional morphometric evidence, but validated sex-estimation equations and accuracy metrics require access to individual-level data and independent testing.

## Data Availability

Data will be made available and shared upon request, by the authors.

## Declarations

## Ethics approval and consent to participate

The study received approval from the Paramedical Research Committee, College of Paramedical Sciences, Teerthanker Mahaveer University. According to the committee guidelines, studies involving routine diagnostic laboratory examinations were exempted from the formal ethical consideration.

## Availability of data and materials

Data will be made available and shared upon request, by the authors.

## Author contributions

Harsh Kumar Singh performed all the experiments and prepared the manuscript. Dr. Raushan Kumar prepared the experimental procedures and carefully analyzed the results. Dr. Swati Dixit reviewed the manuscript and formatted it. thesis research under the guidance of Raushan Kumar.

